# Standardization of drug names in the FDA Adverse Event Reporting System: The DiAna dictionary

**DOI:** 10.1101/2023.06.07.23291076

**Authors:** Michele Fusaroli, Valentina Giunchi, Vera Battini, Stefano Puligheddu, Charles Khouri, Carla Carnovale, Emanuel Raschi, Elisabetta Poluzzi

## Abstract

**Introduction:** The FDA Adverse Event Reporting System (FAERS) receives drug names in various forms, including brand names, active ingredients, abbreviations, and misspellings, which creates challenges in nomenclature standardization. The lack of consensus on standardization strategies and of transparency hampers replicability and accuracy in conducting disproportionality analysis using FAERS data.

**Aim:** We have developed an open-source drug-to-ingredient dictionary called the DiAna dictionary (short for Disproportionality Analysis). Additionally, we have linked the DiAna dictionary to the WHO Anatomic Therapeutic Chemical (ATC) classification system.

**Methods:** We retrieved all drug names reported to the FAERS from 2004 to December 2022. Using existing dictionaries such as RxNorm and string editing techniques, we automatically translated the drug names to active ingredients. Manual revision was performed to correct errors and improve translation accuracy. The resulting DiAna dictionary was linked to the ATC classification, proposing a primary ATC code for each ingredient.

**Results:** We retrieved 18,151,842 reports, with 74,143,411 drug entries. We automatically translated and manually checked the first 14,832 terms, up to terms occurring at least 200 times (96.88% of total drug entries), to 6,282 unique active ingredients. Automatic unchecked translations extend the standardization to 346,854 terms (98.94%). After linking to the ATC classification, the most prominent drug classes in FAERS reports were immunomodulating (37.40%) and nervous system drugs (29.19%).

**Conclusion:** We present the DiAna dictionary as an open-source tool and encourage experts to provide input and feedbacks. Regular updates can improve research quality and promote a common pharmacovigilance toolbox, ultimately advancing safety and improving study interpretability.

**Key points:**

- Drug name standardization impacts signal detection accuracy.
- DiAna dictionary cleanses drugs in FAERS for improved data control.
- DiAna’s transparency and flexibility improves interpretability.

## 1. Introduction

### 1.1. The need for transparency on data preprocessing

Spontaneous reporting systems (SRSs) are public and private services collecting reports of suspected adverse drug reactions in order to timely detect potential issues in drug safety [1,2]. However, these reports are gathered from a variety of regional, national, and manufacturer databases, which have different languages, rules, and forms for data storage. Furthermore, the same SRS may collect reports from different kinds of reporter (e.g., manufacturers, healthcare professionals, lawyers, consumers) and using both paper and electronic forms. This heterogeneity leads to highly variable content, particularly in free text fields which may contain misspellings, out-of-context information, and the use of different lexicons. Furthermore, reports may be incomplete, due to the spontaneous nature of the reporting, or duplicates, due to multiple reporting and system errors. For these reasons, an extensive cleaning procedure is needed prior to any kind of statistical analysis.

The United States (US) Food and Drug Administration (FDA) Adverse Event Reporting System (FAERS) is one of the most used SRS in signal detection, because of its free access and its large catchment area mirroring the entire world (even if with a large representativity for the US). There are two ways the FDA provides free access to the FAERS data, with notable differences.

First, already pre-processed data through an online public dashboard [3], developed for transparency reasons and the promotion of higher quality reporting. However, this tool is obtained from spontaneous reports through a cleaning procedure that is, allegedly, both undisclosed and partial, for example because it does not attempt any duplicates detection and it does not provide all the information reported in individual case safety reports (ICSRs). Therefore, it is not suitable for research purpose, namely complex analyses beyond simple exploratory analyses.

Second, raw quarterly data (both in ASCII and XML format) [4] that allow to knowingly perform and document the entire pre-processing procedure. This cleaning and normalization procedure requires the researchers a conspicuous effort and faces them with the need to make multiple operative choices: for example how to deal with duplicates, how to deal with dates that up to 2012 were completed automatically when partial, how to deal with unclear entries (e.g., in 2019 “RN” was often recorded as a reporter type; while this entry may refer to registered nurses, it is not documented in the “readme” file of the FAERS).

These choices are seldom driven by objectivity alone and must be considered both in the design and the interpretation, because the same database cleaned with different procedures may give different results [5]. The subsequent lack of replicability heavily impacts on the credibility of SRSs, already diminished due to their inherent bias, which hinders any interpretation of disproportionality analysis beyond the generation of hypotheses [6].

To sum up, SRSs’ data are extremely raw and heterogeneous, necessitating a meticulous cleaning process guided by clinical and pharmacological reasoning before conducting any analysis [7]. Throughout each stage of this process, it is crucial to uphold collaboration among multiple professionals and maintain an understanding of the data collection features and the relevant underlying theory for the phenomenon under investigation [8]. Additionally, it is of utmost importance, for researchers, not only to be in control and knowledgeable of the pre-processing procedure, but also to be transparent about their operational choices, allowing for external assessment and interpretation.

### 1.2. Drug nomenclature issues

Among the information requiring standardization prior to case retrieval and analysis, a particular effort should be focused on the standardization of drug names, which in the FAERS are collected as verbatim (free) text. A drug may be recorded in the FAERS using either the brand name or the active ingredients, with the international nonproprietary term (INN, defined by the WHO) or the United States adopted name (USAN, defined by the USAN council), with the full name or abbreviations. Misspellings might easily occur, and the drug name is sometimes followed by dose, route, and formulation details.

An objective standardization of these entries is unattainable: for example, the same brand name may refer to different compositions in different countries, or two brand names may be just one letter apart thus being extremely susceptible to misspellings. The inconsistencies that derive from the multiple operative options and researcher’s personal choices can affect case-retrieval and impair replicability among studies. Nonetheless, no consensus on the best operative procedures has been achieved, and already-published analyses using SRSs’ data are rarely transparent on the cleaning choices adopted, lacking documentation on whether and how the FAERS was prepared for statistical analyses. The FAERS system itself, which does have a formal dictionary through which it cleans the data for the public dashboard, does not make it publicly available. Among the 17 studies conducted using the FAERS quarterly data and published in February 2023 (accessed on PubMed on March 15, 2023), ten did not state any drug standardization process, six used an automatic translation via dictionaries, one performed a manual translation but did not make it publicly available (Table S1). This lack of transparency also affects some free ready-to-use pharmacovigilance databases which provide already pre-processed FAERS data [9,10]. Moreover, these databases standardize drugs only according to US dictionaries of drug names (e.g., RX-norm, orange book), thus failing to identify foreign drug names, misspellings, and other free text issues that were described above. The WHODrug Global is a dictionary that compiles extensive drug information from across the globe [11]. However, it does not consider the challenges posed by free text issues, and it is not available as an open-source resource. Other tools attempt to edit drug names using an automatic translation of potential misspellings, which may result in mistranslations due to the existence of similar drug names referring to formulations with different active ingredients [12]. For this reason, already in 2015 Wong et al. produced a manually revised translation of the LAERS drug names (the FAERS system up to 2012), with a transparent explanation of the operative choices [13]. Still, their translation was not publicly available and did not came into use.

### 1.3. Aim of the study

In this work, we follow Wong et al. efforts and extend their work to consider previously unattended nomenclature issues. We propose an open-source drug name-to-ingredient dictionary for standardizing the FAERS updated to December 2022 together with a transparent report of the data cleaning protocol to identify and resolve drug nomenclature issues. This pharmacovigilance tool, that we define as DiAna (Disproportionality Analyses) dictionary, with its linkage to the Anatomic Therapeutic Chemical (ATC) classification, aims to support pharmacovigilance researchers towards a greater control on the FAERS data, a higher replicability and accuracy of disproportionality analyses, and a more appropriate interpretation of their results.

## 2. Methods

### 2.1. The FAERS database

We downloaded FAERS Quarterly Data (trimestral) Extract Files[4] in ASCII format from 04Q1 to 22Q4. These files are composed of five tables linked through a primary key (“primaryid”) identifying a specific version of a report: DEMO (demographic and administrative information), DRUG (information on reported medications), REAC (adverse events), OUTC (outcomes), RPSR (report sources).

DRUG is also linked through “primaryid” and a secondary key (“drug_seq”), identifying a specific medication within a report, to other two tables: THER (start dates and end dates for the reported medications), INDI (indications for using the reported medications).

### 2.2. Automatic set up of the dictionary

We combined all DRUG quarters into one database. We focused on three columns used to identify the medicinal product:

- Drugname, recording the name of the medicinal product.
- Prod_ai, recording the product’s active ingredients, when available.
- Val_vbm, recording whether the source of drugname was a validated trade name (value = 1) or a verbatim name (value = 2).

Since our aim was the translation to active ingredients, we did not consider the column “val_vbm”. We instead retrieved all the unique terms from the other two columns (i.e., Prod_ai and Drugname), lowered upper cases, and removed multiple spaces, leading and trailing spaces and punctuation, and spaces between parentheses and included text. We merged the unique terms with RX-norm and WHO-ATC substances to create a dictionary with automatic translations to active ingredients. The merging process was also repeated after several rounds of text editing, during which we removed leading or trailing spaces and specific terms or symbols such as chirality indicators (e.g., “+”, “-”, “d”, “s”) and text between brackets or caret symbols.

### 2.3. Manual revision

We then manually revised all the automatic translations starting with the most frequently reported ones up to the terms recorded in a minimum of 200 reports (and beyond, ongoing). We started translating drug names to active ingredients included in the 2023 update of the ATC classification of the WHO, integrating it whenever we met new active ingredients. Hand search of foreign drug names was performed using online databases (e.g., DrugBank.com [14] and Drugs.com [15]), manufacturer websites, and websites storing information from foreign package labels (e.g., Kusuri-no-Shiori –drug information sheets– from the Japanese regulatory agency, accessed at https://www.rad-ar.or.jp/siori/english/).

### 2.4. Nomenclature issues

Multiple issues were identified in the process of translating drug names, including brand names and abbreviations) to active ingredients (e.g., “Zantac” was translated into “raniditine”):

- A drug may include multiple ingredients. We translated the drug to all its ingredients and ordered them alphabetically, separating them by a semicolon. For example, “Entresto” was translated into “sacubitril;valsartan”.
- The spelling of an active ingredient can be different between the United States Adopted Name (USAN) and the International Nonproprietary Name (INN): for example, acetaminophen (USAN) = paracetamol (INN); amphetamine (USAN) = amfetamine (INN); dimethicone/simethicone (USAN) = simeticone (INN); cysteamine (USAN) = mercaptamine (INN). We gave preference to the INN.
- The active ingredient may be recorded in languages different from English (e.g., acide folique). We translated everything to the English INN.
- Typing mistakes can occur (e.g., “zoplicone” instead of “zopiclone”; “Diavan®” instead of “Diovan®”). We manually fixed the mistakes taking into account the INN.
- The same drug name may contain different ingredients in different countries (e.g., Gaster® contains famotidine in the US, omeprazole in Japan, cromoglicic acid in Italy; Previscan® contains fluindione in the US, pentoxyfilline in Italy and Spain; Furix® contains furosemide in the US, cefuroxime in India). In these cases, we translated the brand name to the active ingredient contained in the US packaging, assuming that US is more represented in the FAERS rather than other countries. When the brand name of interest was not sold in the US, we checked the most reported country in the FAERS for that specific brand name.
- The drug name may be missing or underspecified:

`o` When there was no medication, we translated the drug name field to “no medication”.
`o` When the medication was unspecified, we translated the drug name field as “unspecified”.
`o` Unspecified drug-class terms were translated to the most specific term possible (e.g., “water pills” as “diuretics, unspecified”, “antihypertensives” as “antihypertensives, unspecified”).
- We specified when drug (or placebo) consumption occurred in a clinical trial (such as when blinded was specified, or when the investigational name was used –e.g., cc-223 for onatasertib) translating the drug name as “active ingredient, trial”, “placebo, trial”, or “unspecified, trial”.
- Additionally, we decided to standardize terms other than drugs to broader categories, since specific details are seldom provided: minerals (e.g., calcium), vitamins (e.g., vitamin b5, independent of the route of administration; vitamin b12, independent of the form–e.g., cyanocobalamin, mecobalamin–), devices (e.g., intrauterine contraceptive device), vaccines (e.g., COVID-19 vaccine), and phytotherapies (e.g., plantago spp).
- If a drug name was reported followed by a non-coherent active ingredient in square brackets, we assumed that an error was made during the compilation by the pharmacovigilance expert. In this case, we translated the entry based on the drug name alone, considering incorrect the active ingredient listed in square brackets.

The whole list of standardized names is available in the open-source repository [https://osf.io/zqu89/?view_only=237d052047c142cabd5d8ea2e765efc6].

### 2.5. Linkage to the ATC classification

Furthermore, we performed data-linkage between the DiAna dictionary and the hierarchical ATC classification, which was downloaded from the WHO Collaborating Centre for Drug Statistics and Methodology website [16] using the R package “rvest”. Since this classification is mainly a tool for drug utilization research, the same active ingredient may be given more than one ATC code if it is available in multiple strengths or routes of administration with clearly different therapeutic uses [17]. We linked the final list of individual active ingredients from our translation with the ATC classification, manually integrating for different choices in the nomenclature (e.g., “vitamin b9”–folic acid– to B03BB01), for classes of drugs (e.g., “antihypertensives, unspecified” to C02), and for drugs recorded in the ATC only in combination (glecaprevir and pibrentasvir both to J05AP57). We have here linked each active ingredient to all its ATC codes (most of the time it is not possible to discriminate between the different ATC codes based only on the drug name) but, since sometimes it is important to count each ingredient only once, we also proposed a unique primary ATC code for each ingredient. To this end, we prioritized the first level in the following order (“H”,“J”,“P”,“L”,“M”,“N”,“C”,“G”,“R”,“B”,“D”,“A”,“S”,“V”). We furtherly moved vitamin c from G01AD03 to A11GA01, sex hormones having both a genitourinal and an immunomodulating code to the genitourinal, and sodium and calcium chloride to the alimentary ATC code instead of the blood-related. This data linkage, while not useful to identify specific formulations, may be used to define the drugs of interest or for visualization purposes in the implementation of disproportionality analyses.

## 3. Results

We downloaded the FAERS quarterly data up to 22Q4 and retrieved 18,151,842 ICSRs, for a total of 74,143,411 drug entries (92.81% allegedly recorded using a validated trade name) and 955,778 unique drugname and prod_ai terms (see Figure 1). After the initial formatting, we reduced them to 793,274 unique entries. We automatically translated 346,854 terms (98.94% of total drug entries) and manually checked the first 14,832 terms (96.88%) up to 174 occurrences (<0.00015%, ongoing).

**Figure 1.**
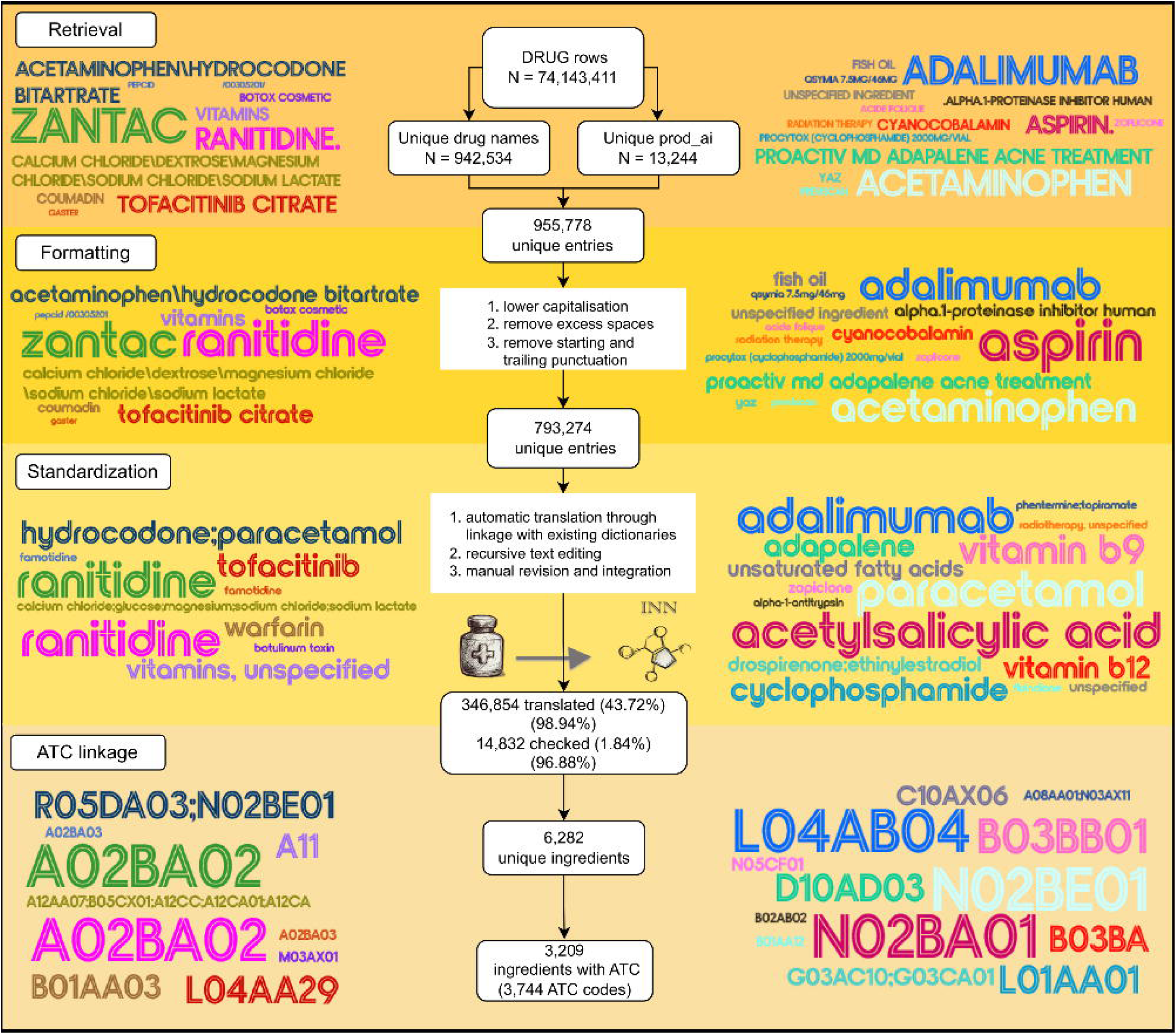
Translation pipeline. Flowchart showing the procedure to translate drug names to active ingredients. Some examples of the processing of entries are provided in the background. The color remains constant across the steps, and within each step the dimension is proportional to the number of occurrences.

A total of 6,282 unique ingredients were included in the DiAna dictionary, of which 3,209 were linked to the ATC classification. The most common primary ATC classes in the FAERS, after translation with the DiAna dictionary, were immunomodulating (reported in 37.40% FAERS reports), nervous system (29.19%), alimentary tract (25.18%), and cardiovascular agents (20.17%) (see Figure 2). Most frequently reported medicinal products were paracetamol (5.45%), acetylsalicylic acid (4.62%), adalimumab (3.81%), etanercept (3.35%), levothyroxine (3.17%), ranitidine (3.13%) (see Table 1). When compared with the untranslated formatted FAERS and with the FAERS translated according to RxNorm, the translation based on DiAna dictionary showed clear advantages in case retrieval (98.94% of total drug entries against 76.32% by RxNorm). Among the most reported medicinal products, DiAna allowed to retrieve more cases than RxNorm, from a ratio of 1.01 for etanercept (638,427 vs 632,130), to a ratio of 8.55 for ranitidine (69,883 vs 597,604). Due to differences in nomenclature some ratios were not calculated (e.g., paracetamol is translated to acetaminophen and acetylsalicylic acid to aspirin by RxNorm). For some drugs the added value of DiAna translation for case retrieval was extremely high: for example, rimegepant (ratio = 277.91; 6,392 vs 23), adapalene (122.60; 174,711 vs 1,425), drospirenone (108.49; 86,356 vs 796), and umeclidinium (105.66; 45,751 vs 433; not shown in the table).

**Table 1.**
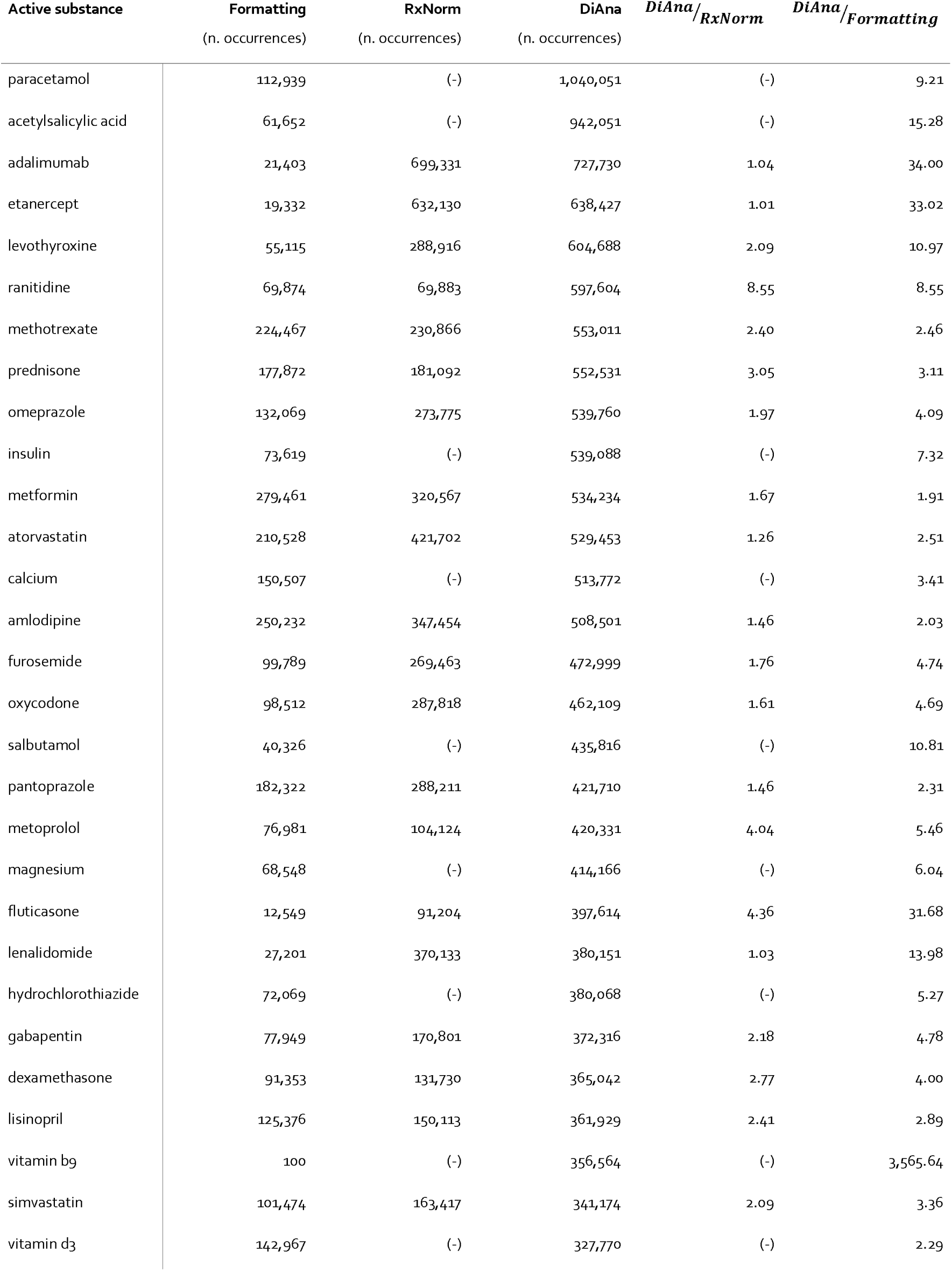
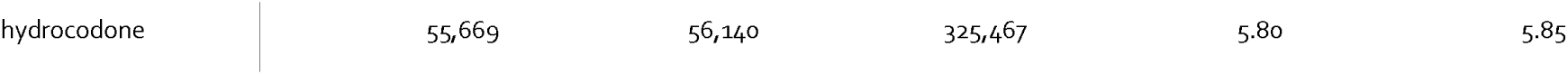
Performance of DiAna translation. Drugs most frequently reported in the FAERS, after DiAna translation, relative to simple formatting and to the merging with RxNorm. The number of occurrences in the three translations are reported together with the ratio of occurrences between DiAna and the others. In some cases, differences in the nomenclature resulted in empty cells.

**Figure 2.**
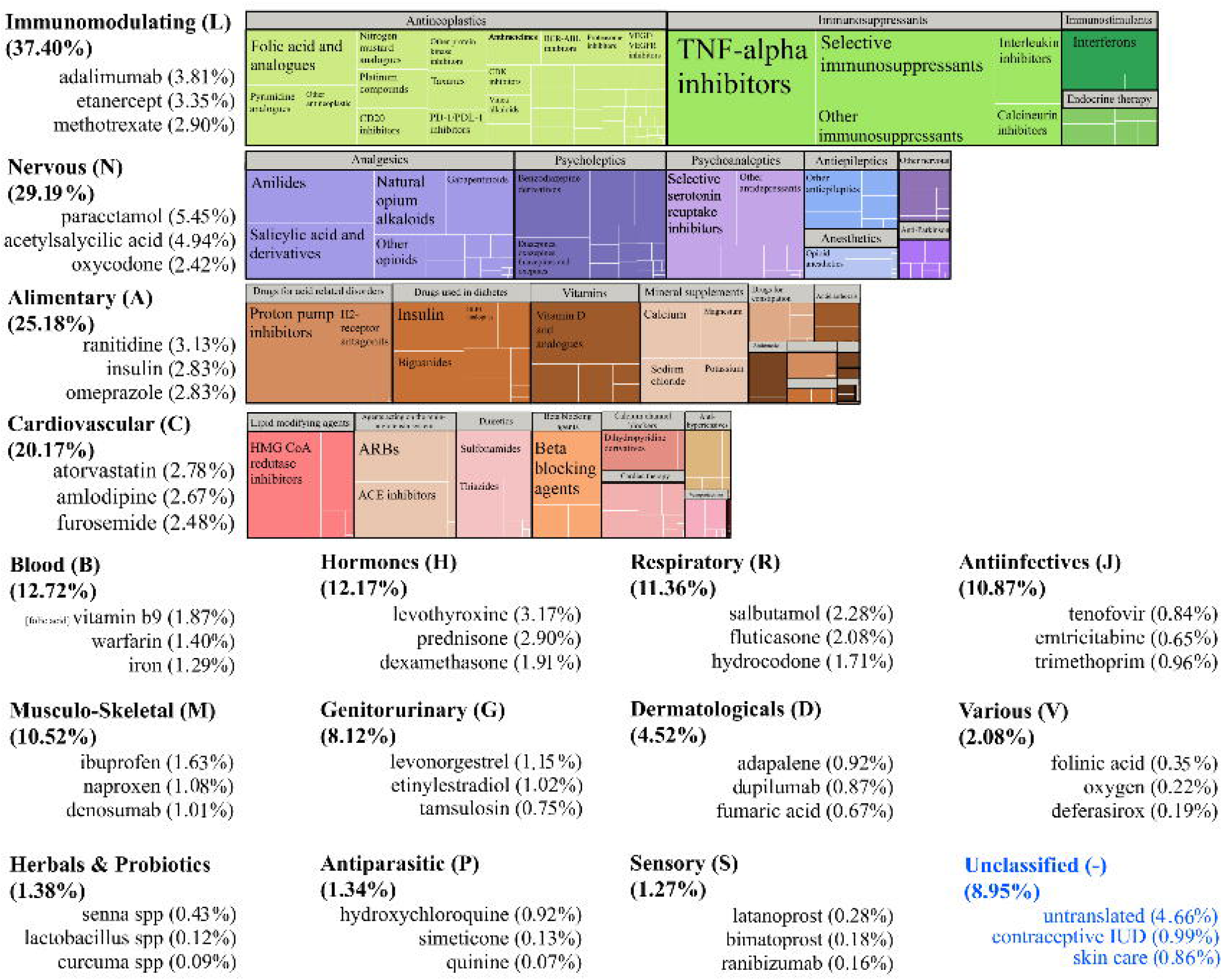
Distribution of medicinal products in the FAERS. Drugs most frequently reported in the FAERS, after translation, according to ATC class. Each step is a first level, starting from the most reported one. Within each level, a tree map shows how ATC levels 2 and 4 are reported in FAERS reports. The 3 most reported active ingredients of each 1st level are also shown.

The translation also took account information about placebo and experiments, thus identifying 50,967 reports as generated within trials (0.28%).

## 4. Discussion

### 4.1. The DiAna Dictionary

The sensitivity of case retrieval and the relevant disproportionality analysis results may vary depending on the drug cleaning procedures used in SRSs. Disproportionality analysis is mostly performed on public dashboards or other analytical tools with no access to underlying data, ready-to-use databases with partial or non-transparent translation, or individually cured undisclosed databases. While these tools provide easy access to disproportionality analysis, they also pose a risk of inappropriate analyses and interpretation due to users’ unawareness on the nature of data. Common drug translation procedures involve automatic linkage to existing dictionaries (offering only partial translation) and automatic algorithms dealing with misspellings (potentially introducing errors). To address these concerns, a dictionary for drug name-to-ingredient translation was developed through an automatic procedure that was manually checked and extended. This dictionary, called DiAna dictionary, required a time-consuming effort and is made available opensource for everyone to use it and propose changes. The use of the DiAna dictionary will allow the pharmacovigilance community to agree on the best possible translation. A greater control on data cleaning will result in improved replicability and accuracy of signals, and more conscious and appropriate interpretation of results, with relevant benefit for the scientific community.

### 4.2. Better retrieval for higher sensitivity

We were able to translate 98.94% of total drug entries to 6,282 unique active ingredients using the DiAna dictionary, compared to 76.32% using only RxNorm. When considering unique drug entries, we translated 346,854 terms over 793,274 (43.72%). We manually checked the first 14,832 terms (up to 174 occurrences), which were responsible for the translation of 96.88% of total drug entries. We believe that this is a good starting point to share our work with the pharmacovigilance community and enable more participative use and development of the DiAna dictionary. In contrast to the previous work by Wong et al.[13], made on the FAERS up to 2012, we made our dictionary (up to 2022) open source. We chose to design the translation so that a new column is produced with only active ingredients, while keeping the original verbatim text in a separate column for more in-depth analyses. We have also decided not to translate to salts as this is rarely taken into account in disproportionality analysis and can lead to confusion about whether the same ingredient with unspecified salt should be considered among cases or non-cases. Instead, we have included the linkage to the ATC classification, and provided translation also to higher ATC classes such as “antihypertensives, unspecified”, as this information can be important for adjusting the analysis and assessing individual cases.

The DiAna dictionary translates a higher proportion of the database, enabling a higher sensitivity in case retrieval, and a higher number of identified cases. This results in better specificity in the definition of non-cases and higher accuracy in signal detection. This means earlier and clearer signals, as in some specific products the number of reports retrieved significantly increased. For example, for rimegepant the DiAna dictionary identifies 278 times more reports than RxNorm alone.

In addition to identifying active ingredients, the drug name information enabled us to identify reports derived from clinical trials (0.28% of total reports), as they recorded placebo, blinding, or drug codes. This information can help researchers exclude evidence already taken into account in other steps of drug safety characterization from the disproportionality analysis.

Finally, the linkage between the DiAna dictionary and the ATC classification can help in the retrieval of drug classes and in visualization. The information on the distribution of drug classes in the database is particularly useful for the design of future disproportionality analyses, as it provides insight into the representativeness of the population chosen as comparison. Over one-third of the database consists of reports documenting the utilisation of immunomodulating drugs. Recent observations, specifically in the context of the extensive rollout of COVID-19 vaccines, have reignited the attention to the possibility that this uneven distribution of drugs in the SRS may lead to masking/cloaking bias, hiding disproportionality signals [18].

The DiAna dictionary and its linkage to the ATC classification are freely available online for everyone to use (https://osf.io/zqu89/view_only=237d052047c142cabd5d8ea2e765efc6), and can be corrected and expanded by experts in the field. Changes can be proposed in the GitHub repository (https://github.com/fusarolimichele/DiAna) under the issue DiAna dictionary and will be periodically validated and integrated into the existing dictionary. This collaborative effort will improve the quality and reproducibility of pharmacovigilance research. The dictionary can be downloaded in Excel and csv formats and can be imported into any data management software, such as R, to automatically translate drug names to active ingredients before conducting analyses. Users can also easily modify the translation of specific terms for their analyses, which is not possible with ready-to-use FAERS databases.

### 4.3. Limitations, Strengths, and Further Goals

The DiAna dictionary is not designed as a static dictionary: it will require ongoing efforts to keep up with new drugs and terms. We are recursively extending our translation to reach and maintain a full checked translation of any entry with over 100 occurrences. Users of the DiAna dictionary should be aware of this limitation (which is even more impairing in other disproportionality tools), especially with less frequent terms that may not be included in the dictionary. It’s recommended that before any research on a specific drug, inherent terms are checked in the dictionary and any new translations are shared to integrate into the DiAna dictionary for everyone to benefit. The translation will plausibly never be complete, since some terms are not easily translated (e.g., “chinese food”) and many choices are partly subjective. However, these choices can be defined in agreement with the entire pharmacovigilance community.

The translation of ambiguous terms was also noted as a challenge, especially with over-the-counter cold, cough, and flu agents (multiple ingredients changing over the years). When we were not certain, we used the higher-level term (e.g., “cough preparations, unspecified”). The lack of expertise in supplements and phytotherapies may have resulted in the dictionary being excessively generic (for example referring to plantago spp instead of individual species, and to covid 19 vaccines instead of specific types), and it could benefit from expert refinement for higher specificity and coverage of entries provided to other spontaneous report databases (CAERS and VAERS are more appropriate to investigate the safety profile of these medicinal products).

Since lack of completeness is a known problem in spontaneous reports, and other information is not always available, we implemented sharp-cut operative choices to retrieve active ingredients based only on the drug name. The use of additional columns such as country, year of occurrence, dose, indication, and route of administration, could help discriminate between mistranslations when the same drug name may be translated to multiple active ingredients. Moreover, information from the drug name column could be used to impute information into other columns. For example, “nizoral a-d” is translated to ketoconazole and refers specifically to an anti-dandruff shampoo (i.e., the indication, formulation, and route of administration could be imputed if missing), while “hypersal” refers to a sodium chloride nebulizer solution, and “jinarc” refers to a formulation of tolvaptan specifically indicated for autosomal dominant polycystic disease. By incorporating a drug name-to-product translation feature, for example referring to the WHO Drug Global or to the Identification of Medicinal Products (IDMP), we could streamline the process of imputation of structured fields using free text, thereby enhancing the value of the DiAna dictionary.

Linking INN names to ATC codes was a complex task due to the existence of combination products (e.g., glecaprevir and pibrentasvir), medicinal products with ingredients that do not have an ATC code yet, and experimental substances which are missing even the INN. The linkage will be annually updated according to changes in the ATC classification to preserve its utility.

## 5. Conclusion

We offer the DiAna dictionary as open-source tool for the pharmacovigilance community to standardize drug names in the FAERS database. Its public accessibility, transparency, and flexibility provide a foundation for ongoing improvement and refinement through input from experts in the field. With periodic updates, this open-source project can drive a common effort towards a more transparent and cleaner shared FAERS database, leading to more replicable and higher quality research in pharmacovigilance. Ultimately, by sharing and mutually enriching our knowledge, we can develop a common pharmacovigilance toolbox that advances safety and improves the accuracy, replicability and reliability of pharmacovigilance studies.

## Data Availability

Availability of data and material: The dictionary and the linkage to the ATC classification are available at https://osf.io/zqu89/?view_only=237d052047c142cabd5d8ea2e765efc6. Code availability: All the processing, analyses, and visualization were obtained through R-software (version 4.2.1). The code for using the dictionary in the cleaning of the FAERS database is available at https://github.com/fusarolimichele/DiAna.

## Acknowledgements

The results, discussion, and conclusions of the study are those of the authors alone and in no way represent the position of the WHO Collaborating Centre for Drug Statistics Methodology on this subject.

MF is enrolled in the PhD in General Medical and Services Sciences, Università degli studi di Bologna, which supports their fellowship. V.G. is supported by EU funds (Programma Operativo Nazionale Italian funds for green and innovative research based on European Structural and Investment Funds). VB is enrolled in the PhD in Experimental and Clinical Pharmacological Sciences, Università degli Studi di Milano, which supports her fellowship. We thank Chiara Ballarin, Margherita Bonaiuti, and Laura Pierantozzi (University of Bologna) for the help in the manual revision of drug names standardisation.

