## Supplemental Table 1 for "Standardization of drug names in the FDA Adverse Event Reporting System: The DiAna dictionary"

### Running Title: The DiAna dictionary for drug standardization

**Authors:** Michele Fusaroli^1^*, Valentina Giunchi^1^*, Vera Battini^2^, Stefano Puligheddu^1^, Charles Khouri^3,4^, Carla Carnovale^2^, Emanuel Raschi^1^, Elisabetta Poluzzi^1^

**Affiliations**:

*^1^Unit of Pharmacology, Department of Medical and Surgical Sciences, University of Bologna, Bologna, Italy.*

*^2^* *Unit of Clinical Pharmacology, Department of Biomedical and Clinical Sciences (DIBIC), ASST Fatebenefratelli-Sacco University Hospital, Università degli Studi di Milano, Milan, Italy.*

*^3^ Grenoble Alpes University Hospital, Pharmacovigilance Unit, Grenoble, France*

*^4^ Univ. Grenoble Alpes, HP2 Laboratory, Inserm U1300, Grenoble, France*

| **pmid** | **access** | **source** | **transparence** |
| --- | --- | --- | --- |
| 36739513 |  | public dashboard |  |
| 36274626 |  | public dashboard |  |
| 36725376 |  | open vigil |  |
| 36797979 |  | not specified |  |
| 36875514 |  | quarterly data | false |
| 35871395 |  | quarterly data | false |
| 36270450 |  | quarterly data | false |
| 36873988 |  | empirical signal software |  |
| 36736956 |  | quarterly data | true |
| 36865915 |  | quarterly data | false |
| 35996166 |  | not specified |  |
| 36835526 |  | not specified |  |
| 36821807 |  | public dashboard |  |
| 35973681 |  | quarterly data | false |
| 36697083 | No |  |  |
| 36896641 |  | open vigil |  |
| 36847276 |  | not specified |  |
| 36909171 |  | quarterly data | false |
| 36852785 |  | open vigil |  |
| 36806781 |  | not specified |  |
| 36794339 | No |  |  |
| 36865057 |  | quarterly data | false |
| 36350532 |  | quarterly data | true |
| 36803342 | No |  |  |
| 35349180 |  | quarterly data | true |
| 36700393 |  | quarterly data | true |
| 36651638 |  | quarterly data | true |
| 36848023 |  | not specified |  |
| 36637688 |  | quarterly data | false |
| 36106653 |  | quarterly data | false |
| 36794347 | No |  |  |
| 36734333 |  | quarterly data | false |
| 36829251 |  | quarterly data | true |
| 36371585 |  | quarterly data | true |
| 36794498 | No |  |  |

**Table S1 – Drug standardization in published disproportionality analysis.** In gray the manuscripts excluded from the final count because not accessible or using alternatives to quarterly data.
